# Metabolic Responses to an Acute Glucose Challenge: The Differential Effects of Eight Weeks of Almond vs. Cracker Consumption in Young Adults

**DOI:** 10.1101/2024.05.19.24307571

**Authors:** Jaapna Dhillon, Saurabh Pandey, John W. Newman, Oliver Fiehn, Rudy M. Ortiz

## Abstract

This study investigated the dynamic responses to an acute glucose challenge following chronic almond versus cracker consumption for 8 weeks (clinicaltrials.gov ID: NCT03084003). Seventy-three young adults (age: 18-19 years, BMI: 18-41 kg/m^2^) participated in an 8-week randomized, controlled, parallel-arm intervention and were randomly assigned to consume either almonds (2 oz/d, n=38) or an isocaloric control snack of graham crackers (325 kcal/d, n=35) daily for 8 weeks. Twenty participants from each group underwent a 2-hour oral glucose tolerance test (oGTT) at the end of the 8-week intervention. Metabolite abundances in the oGTT serum samples were quantified using untargeted metabolomics, and targeted analyses for free PUFAs, total fatty acids, oxylipins, and endocannabinoids. Multivariate, univariate, and chemical enrichment analyses were conducted to identify significant metabolic shifts. Findings exhibit a biphasic lipid response distinguished by higher levels of unsaturated triglycerides in the earlier periods of the oGTT followed by lower levels in the latter period in the almond versus cracker group (p-value<0.05, chemical enrichment analyses). Almond (vs. cracker) consumption was also associated with higher AUC_120 min_ of aminomalonate, and oxylipins (p-value<0.05), but lower AUC_120 min_ of L-cystine, N-acetylmannosamine, and isoheptadecanoic acid (p-value<0.05). Additionally, the Matsuda Index in the almond group correlated with AUC_120 min_ of CE 22:6 (r=- 0.46; p-value<0.05) and 12,13 DiHOME (r=0.45; p-value<0.05). Almond consumption for 8 weeks leads to dynamic, differential shifts in response to an acute glucose challenge, marked by alterations in lipid and amino acid mediators involved in metabolic and physiological pathways.

## INTRODUCTION

The intricate relationship between diet, metabolism, and health has been a focal point of nutritional research for decades. Recent advances in metabolomics and nutrition science have enabled a more detailed understanding of how specific foods influence metabolic pathways and, consequently, health outcomes. However, very few dietary studies have examined the dynamic changes in the metabolome in response to an acute glucose challenge. In one such study, a weight loss intervention resulted in differential changes in oGTT area under the curve (AUC) for 11 metabolites before and after the intervention [1]. Although the glucose AUC reduction did not reach statistical significance, the glucose challenge effectively revealed metabolic patterns associated with metabolic health. For example, increased AUCs of α-ketoglutarate, arachidic acid (C20∶0), and the gut microbe-derived compound, tricarballylic acid, were observed post-intervention [1].

Among various dietary interventions, the consumption of nuts, particularly almonds, has garnered attention due to their unique nutrient composition and potential health benefits [2,3]. Almonds are rich in unsaturated fats, protein, fiber, vitamins, and bioactive compounds, which collectively contribute to their proposed health benefits [2]. In our prior work, almond snacking was found to enhance postprandial glucose regulation [4] and fasting serum metabolomic profiles [5], showcasing distinct advantages over snacking on graham crackers, a low fat carbohydrate rich food. More specifically, almond consumption for 8 weeks led to changes in lipid metabolism and the tricarboxylic acid (TCA) cycle, along with possible shifts in microbial synthesis of amino acids, and the metabolism of amino and nucleotide sugars [5]. Additionally, we noted a correlation between the predicted microbial community metabolic potential (CMP) score for N-acetyl-D-mannosamine and the host serum levels of this metabolite [5].

Building on these findings, the current study employs both untargeted and targeted metabolomic approaches to further investigate the oGTT-induced dynamic changes in serum metabolites and underlying metabolic pathways following 8 weeks of almond or cracker consumption. This study, therefore, contributes to the expanding field of nutritional metabolomics, focusing on how functional foods can modulate cellular metabolism reflected in the changes in the human metabolome in the context of a metabolic challenge. The strength of the current approach is that it examines the acute, dynamic metabolic changes stimulated by an acute, substrate challenge as opposed to the more traditional examination of the static, single-sample changes observed at the end of the study. Understanding the metabolic alterations induced by foods can offer new insights into the role of diet in metabolic health and disease. It also holds the potential to inform dietary guidelines and interventions aimed at improving metabolic outcomes.

## METHODS

The University of California (UC) Merced Institutional Review Board granted approval for all procedures involving human subjects in this study. The study is registered on ClinicalTrials.gov under the registration number NCT03084003.

### Participants

A total of seventy-three young adults (41 women and 32 men), aged 18-19 years with a BMI range of 18-41 kg/m2, were enrolled in an 8-week randomized, controlled, parallel-arm intervention. The study aimed to investigate the impact of almond versus cracker snacking on cardiometabolic, microbiome, and metabolomics outcomes. The eligibility criteria and recruitment criteria have been described in detail previously [4].

### Study design and protocol

Our previous publication [4] provides a detailed description of the primary study design. Sample size calculations for the main analysis were based on glucoregulatory profiles after the 8-week intervention. Randomization assigned participants to either the almond group (n =38) or the cracker group (n =35). The almond group consumed 57 g/d (2 oz) of whole, dry-roasted almonds, while the cracker group served as the isocaloric control and consumed 5 sheets (77.5 g/d) of graham crackers. An oral glucose tolerance test was conducted at the end of the 8 wk intervention according to established protocols [4]. A subset of 20 participants per group were considered for the oGTT analysis. Serum samples were collected prior to and at 15, 30, 60, and 120 min during the oral glucose tolerance test and stored at -80 °C.

### Metabolomics analyses

Metabolomics analyses were conducted using various techniques. Gas chromatography-time-of-flight mass spectrometry (GC-TOF MS), hydrophilic interaction liquid chromatography (HILIC) MS/MS, and charged surface hybrid-reversed phase liquid chromatography electrospray (CSH-ESI) MS/MS were employed for analysis of serum samples as previously described [5]. Quantitative ion peak heights were used to report the data, and known metabolites underwent normalization using the Systematic Error Removal Using Random Forest (SERRF) method [6]. Metabolites with a QC RSD greater than 50% were excluded from subsequent analyses. For HILIC-MS, metabolite intensities were compared to blanks. Metabolites that did not exhibit significant differences between the sample and blank (p-value >0.05 in the Wilcoxon test) and those with a median sample to blank ratio less than 1 were also excluded from further analyses. Quantitative targeted analyses of total alkali-releasable fatty acids, and non-esterified PUFAs, oxylipins, and endocannabinoids were also performed by MS/MS using internal standard methodologies and authentic standards as previously described [5]. For each metabolite, the area under the curve (AUC) over the different time periods of the 120 min-oGTT was calculated by trapezoidal rule integrations.

### Multivariate and univariate analyses of metabolomics data

Partial least squares discriminant analysis (PLS-DA) was conducted in JMP Pro (version 17.0) to identify shifts in metabolites over the oGTT time points (0,15, 30, 60, 120). Before the analysis, missing values were imputed using multivariate imputations [7]. Models were calculated using autoscaling on log10 transformed data followed by nonlinear iterative partial least squares (NIPALS) with leave-one-out cross validation. Variables underwent a pruning process, where initially all variables were considered, then in each subsequent step, only those with variable importance in projection (VIP) scores exceeding 0.8 were retained, resulting in a final selection which optimized the variability in the predictor data set. A repeated measures analysis on the latent variables with timepoint as within-subject factor were also conducted in JMP Pro.

The selected PLS-DA variables were subjected to fuzzy c-means clustering with the Mfuzz package [8] in R to categorize metabolites into groups based on their time-course patterns [9]. For clustering purposes, each metabolite’s z-score was calculated from the average value of all subjects at each time point. Optimal clustering parameters (fuzzifier *m*) for the data were estimated by the Mfuzz package (m=2.43). The clustering process assigned each metabolite a membership probability for each cluster, and metabolites were then assigned to the cluster for which they had the highest membership probability.

For the univariate analyses, a 2-step approach to the analyses was deployed. The first step focused on selecting the identified metabolites that showed a significant overall time effect in a linear, mixed model analysis with time (0, 15, 30, 60, and 120 min) as the factor. The time effect p-values for metabolites were corrected for multiple hypotheses testing using Benjamini-Hochberg correction (false discovery rate (FDR) adjusted p-value). Metabolites that demonstrated a significant (FDR <0.05) time effect were selected for further analyses. The second step was comprised of a linear model analysis for selected metabolite AUCs with snack group as a factor, with analyses adjusted for time 0 values. All data are reported as means and SDs unless otherwise stated. The univariate statistical analyses were performed using R version 4.2.1

### Chemical enrichment analysis

Chemical enrichment analyses were performed using ChemRICH, as described previously [5]. The dataset used for analysis included time 0-adjusted overall snack effect p-values for all annotated metabolites. To determine statistically significant p-values for metabolite clusters, Kolmogorov-Smirnov tests were conducted and adjusted for false discovery rate (FDR).

### Network analysis

To explore differences between the almond and cracker groups within a biochemical and structural context, network analysis was conducted. The parameters for the construction of the network map using MetaMapp [10] and Cytoscape 3.7.2 [11] have been described previously [5]. The quantitative data set was comprised of the overall effect size (Hedge’s g for almond vs cracker AUCs) and time 0-adjusted snack effect p-values. Since this was an exploratory analysis, the p-values were not adjusted for false discovery rate (FDR). The network map only shows metabolites with time 0-adjusted snack effect P <0.05 or those with large effect sizes (Hedge’s g >0.8).

### Correlation analyses

An exploratory aim of the current work was to identify metabolites that correlate with insulin sensitivity as measured by the Matsuda Index (MI) for the almond and cracker groups differentially. Insulin sensitivity has been found to be differentially influenced by almond consumption [4]. PLS-DA was conducted in JMP Pro (version 17.0) to identify optimal metabolite AUCS and indices between almond and cracker groups. Before the analysis, missing values were imputed using the multivariate normal imputation method in JMP Pro [12]. Models were calculated using autoscaling on log10 transformed data followed by NIPALS with leave-one-out cross validation. The model was pruned until optimization, and variables with VIP >0.8 were selected for further analyses. Variables with loading values for latent variable 1 that were at least 1.5 SD different to the mean loading values were selected for correlation analysis. After verifying data normality by Shapiro-Wilk’s test and visual observation of the normal quantile plots, Pearson’s correlations were computed for those variables. To assess the statistical significance of the correlation coefficients between the almond and cracker groups, Fisher’s r to z transformation was employed.

## RESULTS

### Progressive shift in metabolites in response to a glucose challenge

The PLS-DA analysis demonstrates a progressive shift in metabolites in response to a glucose challenge (**Figure 1**) with latent variable 1 values for time 0, 15, 30, 60, and 120 min being different from each other (p-value<0.05 for all comparisons except two: 0 vs. 60 min; p =0.07; 0 vs. 15 min; p >0.05). The cluster analysis revealed four clusters with distinct time course patterns (**Figure 2**). Cluster 1 exhibits a consistent decline in metabolite z-scores from time 0 to 120 min, which implies a sustained decrease in response to a glucose challenge. Cluster 2 shows an overall increasing trend in metabolite z-scores from time 0 to 120 min, with a shallower slope between 60 and 120 min, suggesting these metabolites may be involved in later metabolic responses to glucose ingestion. Cluster 3 demonstrates a ‘V’ shaped pattern with an initial decline at 15 min, followed by a steady increase to 120 min, indicating a strong decline in those metabolite z-scores in response to glucose that gradually increases over time. Cluster 4 displays a fluctuating pattern with an initial increase in metabolite z-scores over the first 30 min followed by a decrease to 120 minutes.

**Figure 1.**
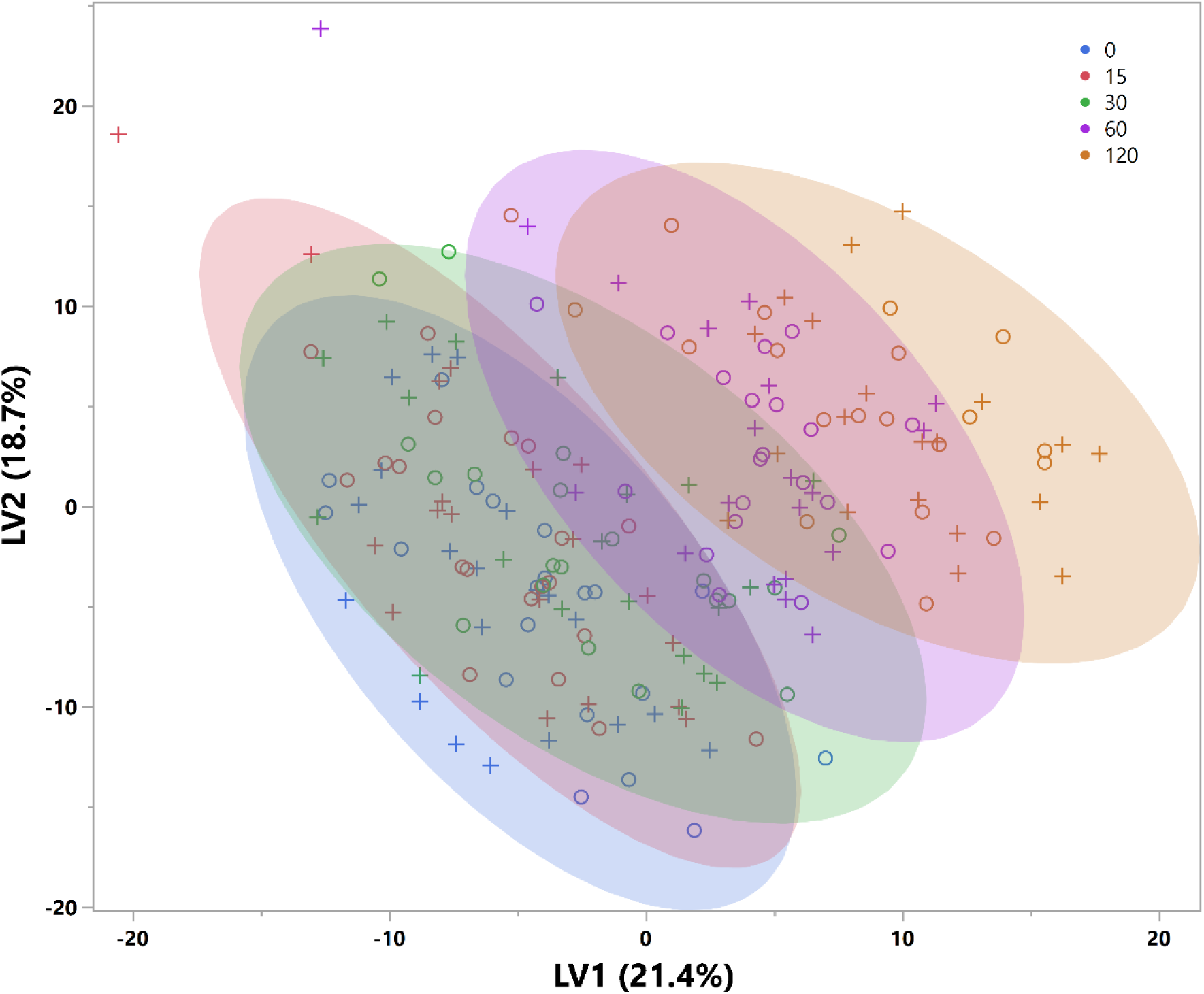
Plot of latent variable 1 versus 2 obtained from the PLS-DA analysis conducted on the metabolites to differentiate among the time points of the oGTT. circle = almond; plus sign = cracker.

**Figure 2.**
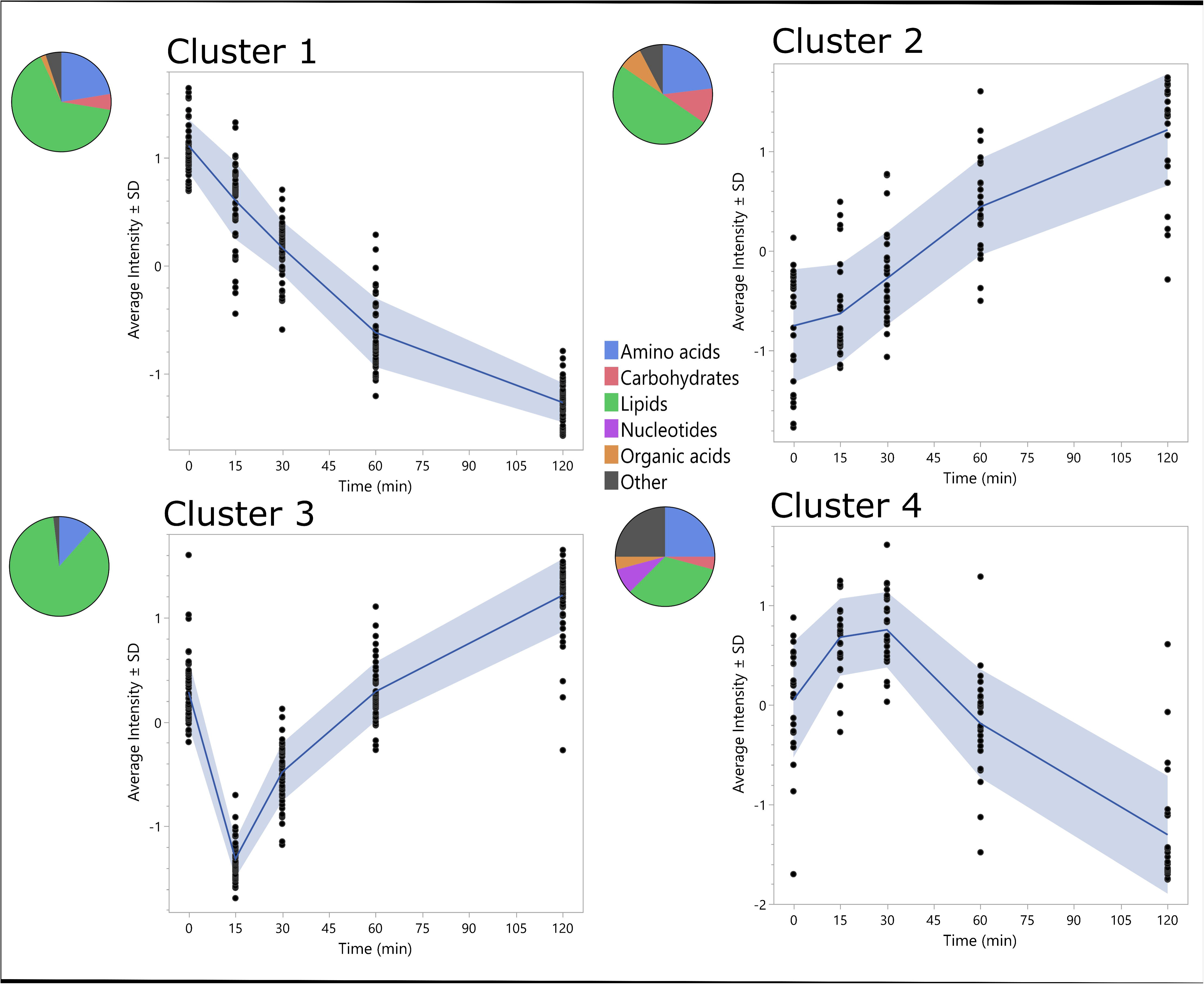
Temporal metabolite patterns in response to an oGTT identified by fuzzy c-means cluster analysis. Graphs show four patterns (clusters) of metabolite trajectories (A–D). The blue line depicts the mean z-score of metabolite trajectories. The colored pie charts depict the breakdown of metabolite type. Similar metabolites are grouped into one category, for example, amino acid derivates, peptides, and amino acids, are all categorized as amino acids, and nucleic acids and nucleotides are categorized as nucleotides.

A total of 268 metabolites had significant (FDR <0.05) overall time effect during the oGTT. The metabolite AUC_120min_ values that had time 0-adjusted snack p-values <0.05 are shown in **Table 1**. At an α =0.05 relative to the graham cracker snack group, almond consumption resulted in higher AUCs of 1/2-OG (39% group difference), Asp-Phe (31%), aminomalonate (24%), arachidonic acid (AA) (15%), 9,10-e-DiHO (6%), 12(13)-EpOME (2%), and lower AUCs of N-acetylmannosamine (63%), isoheptadecanoic acid (28%), N-methylvaline (24%), L-cystine (10%), 15-HETE (3%), 9,10-DiHODE (3%), and 11-HETE (1%). The time course of these metabolites is depicted in **Figure 3**.

**Figure 3.**
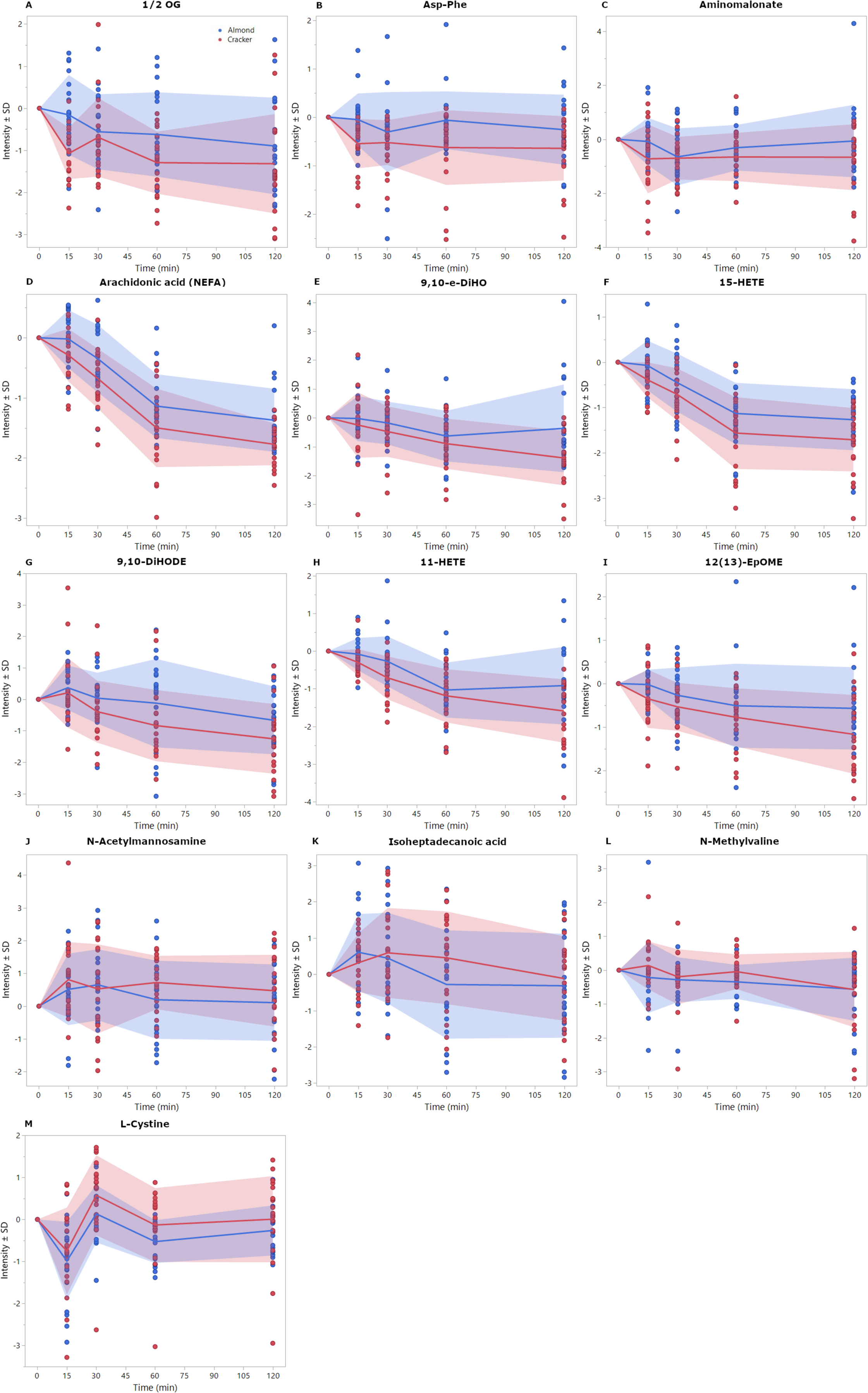
Time 0-adjusted mean intensities ± standard deviation of metabolic trajectories (over 120 min) for selected metabolites that indicated significant (p-value <0.05) time 0-adjusted group effect for AUCs (Table 1). Data are JN transformed.

**Table 1:** Selected metabolites derived from the oGTT AUC_120min_ calculations in the almond and cracker groups after the 8-week intervention.

| Metabolites<sup>a</sup> | Almond | Cracker | Group percent difference | Snack effect P-value (time 0 adjusted)<sup>c</sup> |
| --- | --- | --- | --- | --- |
| *Higher in Almond Group* |  |  |  |  |
| 1/2-OG (x 10<sup>3</sup>) | 40.1 ± 13.4<sup>b</sup> | 35.0 ± 16.0 | 39.2 | 0.002 |
| Asp-Phe (x 10<sup>4</sup>) | 32.0 ± 9.3 | 29.2 ± 8.4 | 31.0 | 0.009 |
| Aminomalonate (x 10<sup>5</sup>) | 24.4 ± 7.3 | 20.3 ± 4.0 | 23.5 | 0.018 |
| AA NEFA (x 10<sup>3</sup>) | 16.1 ± 6.4 | 15.2 ± 4.7 | 15.1 | 0.049 |
| 9, 10-e-DiHO (x 10<sup>2</sup>) | 6.5 ± 2.8 | 5.7 ± 2.7 | 5.6 | 0.033 |
| 15-HETE (x 10<sup>2</sup>) | 4.3 ± 3.6 | 3.6 ± 3.0 | 3.4 | 0.012 |
| 9,10-DiHODE (x 10) | 4.2 ± 3.0 | 3.6 ± 4.5 | 3.3 | 0.002 |
| 11-HETE (x 10<sup>2</sup>) | 2.3 ± 2.3 | 1.9 ± 1.9 | 1.4 | 0.021 |
| 12(13)-EpOME (x 10<sup>2</sup>) | 3.1 ± 4.2 | 2.0 ± 2.2 | 2.3 | 0.021 |
| *Lower in Almond Group* |  |  |  |  |
| N-Acetylmannosamine (x 10<sup>3</sup>) | 64.2 ± 12.3 | 76.0 ± 22.4 | 63.1 | 0.043 |
| Isoheptadecanoic acid (x 10<sup>4</sup>) | 29.1 ± 6.5 | 36.3 ± 1.0 | 28.0 | 0.014 |
| N-Methylvaline (x 10<sup>5</sup>) | 25.3 ± 16.9 | 31.3 ± 19.5 | 24.2 | 0.050 |
| L-Cystine (x 10<sup>5</sup>) | 11.3 ± 0.9 | 12.0 ± 1.7 | 10.3 | 0.023 |

<sup>a</sup> Metabolites were selected based on significant overall time effect, FDR adjusted p-value <0.05
<sup>b</sup> Means ± standard deviation
<sup>c</sup> Linear model analysis with time 0 adjustment
Values in parentheses denote the units of measurement for metabolite intensities. For example, (x 10<sup>3</sup>) indicates intensities in units of 1000. This notation is used to describe large numerical values conveniently.

### MESH class enrichment analysis identifies snack specific early and late period oGTT effects

ChemRICH analysis was conducted on time 0-adjusted AUC values for metabolites over different time intervals. A total of 50 MESH clusters were identified. Over the first 15-minute, and 30-minute periods, there was differential enrichment of diglycerides, unsaturated triglycerides, and lysophosphatidylcholines clusters with the almond group having greater AUC_15 and 30 min_ compared to the cracker group, while the almond group had lower enrichment of AUC _60-120min_ for unsaturated triglycerides, saturated triglycerides, and unsaturated phosphatidylcholines (FDR <0.05). The key metabolites in each of those clusters are depicted in **Table 2**.

**Table 2.**
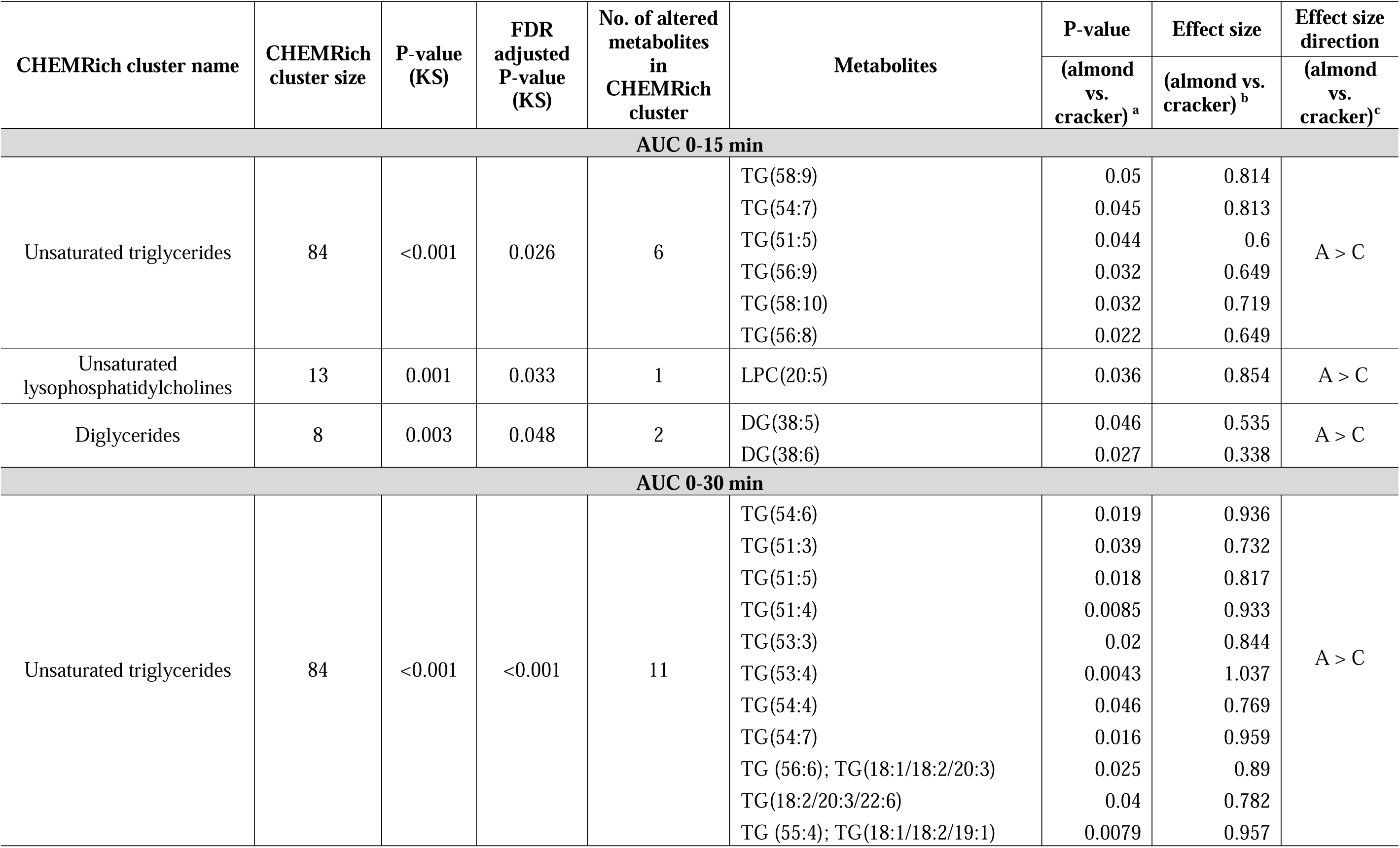
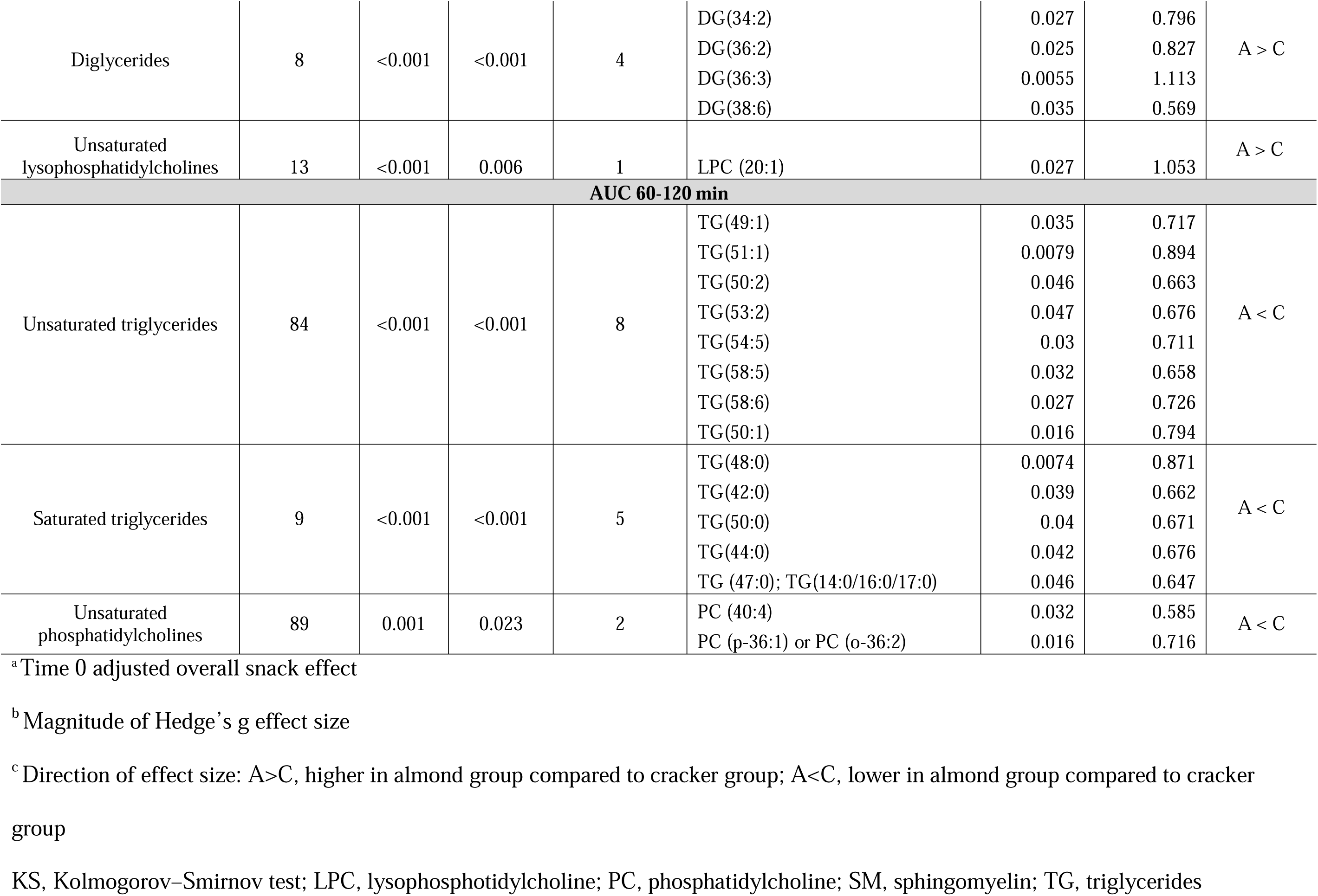
Chemical similarity enrichment analysis results depicting differential AUC clusters of untargeted serum metabolites at the end of the 8-week almond versus cracker intervention

### Amino acid, carbohydrate and lipid metabolism clusters were enriched during the oGTT in response to almond vs. cracker consumption

Enriched AUC_120min_ clusters identified by ChemRich based on metabolic pathway enrichment were largely comprised of metabolites involved in amino acid, carbohydrate, and lipid metabolism. In the network analysis, the metabolites were clustered into groups based on similar structural and biochemical attributes (**Figure 4**).

**Figure 4.**
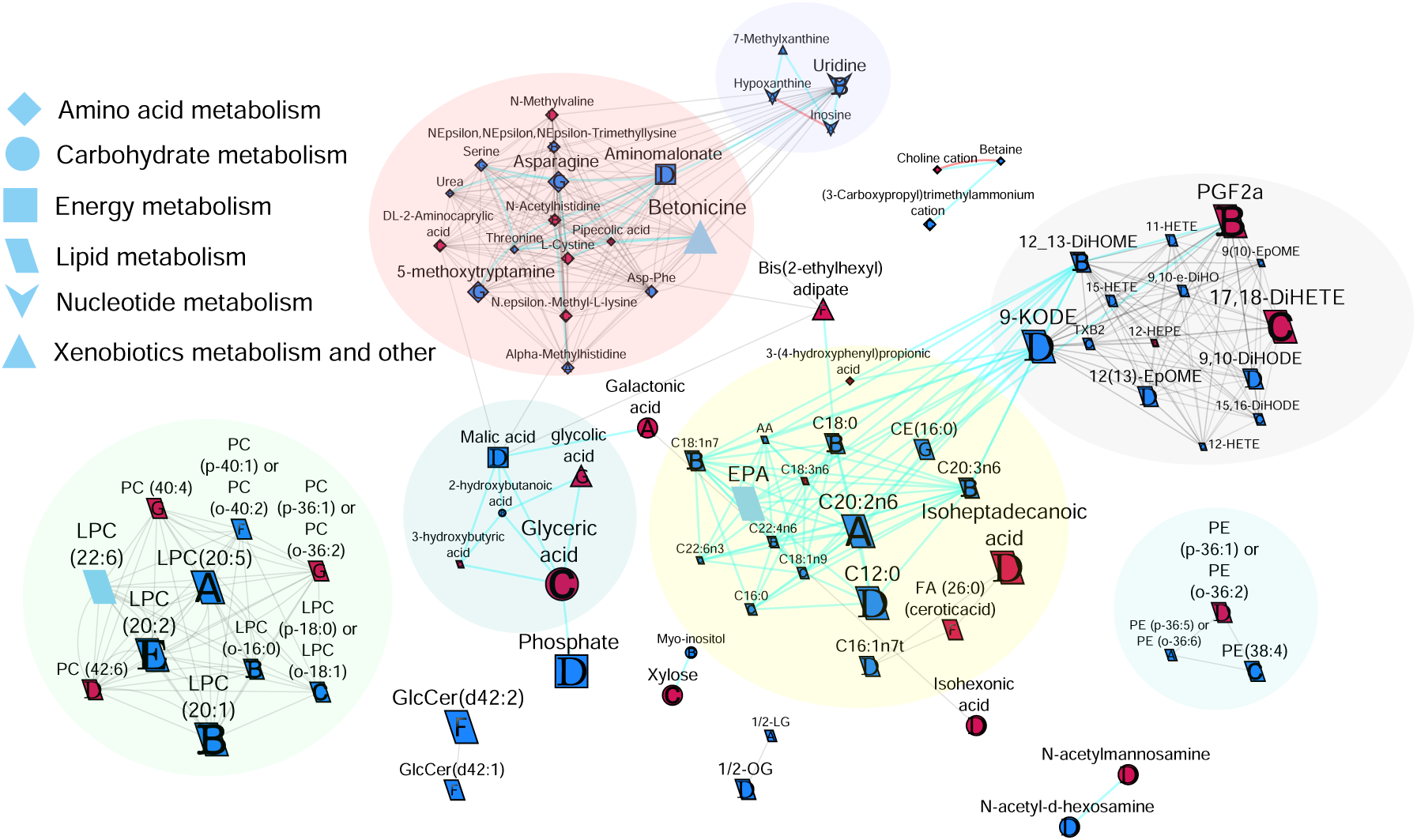
Biochemical network displaying differences between almond and cracker group AUCs over different time periods. Metabolites are connected based on biochemical relationships (orange, KEGG RPAIRS), measured structural similarity (blue, Tanimoto coefficient ≥ 0.7), or manually annotated structural similarity (grey). Metabolite size denotes the effect size (Hedge’s g, almond vs cracker group). Metabolite color represents the direction of the effect size, for example blue represents almond > cracker overall (p-value<0.05) and pink represents almond < cracker overall (p-value<0.05). Letters denote the time period for which significant (p-value<0.05) group effects were observed. For example, “A” represents AUC_0-15 min_ only, “B” represents AUC_0-30 min_ only or in combination with AUC_0-15 min_, “C” represents AUC_0-60 min_ only or in combination with AUC_0-15 min_, AUC_0-30 min_, “D” represents AUC_0-120min_ only or in combination with AUC_0-15 min_, AUC_0-30 min,_ or AUC_0-60 min_, “E” represents AUC_15-30 min_ only, “F” represents AUC_30-60 min_ only, and “G” represents AUC_60-120 min_ only or in combination with AUC_30-60 min_. If metabolites showed significant changes over multiple time periods, only the time period showing the largest effect size was colored. P-values are derived from the time 0-adjusted linear model analysis. Shapes display primary metabolic pathways or structural superclass designations obtained via ClassyFire. Clusters of metabolites are circled. Significant metabolites which did not have KEGG identifiers are included as independent nodes with manually annotated edges within their respective pathway clusters. Diglyceride and triglyceride clusters are not depicted in this network map for clarity. CE, cholesterol ester; LPC, lysophosphotidylcholine; PC, phosphotidylcholine; PE, phosphatidylethanolamine.

### Metabolites involved in amino acid metabolism

Almond vs. cracker consumption had differential effects on several amino acid metabolite AUCs over different time intervals. The almond group had lower AUC_120min_ for L-cystine and N-methylvaline and greater AUC_120min_ for threonine and Asp-Phe in comparison to the cracker group (group effect, p-value <0.05). Over the 60-minute period, the almond group also had lower AUC_60 min_ for N-ε-methyl-L-lysine, DL-2-aminocaprylic acid, choline cation and higher AUC_60 min_ for (3-carboxypropyl)trimethylammonium cation, and betaine compared to the cracker group (group effect, p-value <0.05). Differential group effects for various metabolites were also observed over the 15- and 30-minute periods (**Figure 4**).

### Metabolites involved in carbohydrate metabolism

Almond group had lower AUC_120min_ for N-acetylmannosamine and isohexonic but greater AUC_120 min_ for N-acetyl-D-hexosamine in comparison to the cracker group (group effect, p-value <0.05). Over the 60-minute period, the almond group had lower AUC_60min_ for glyceric acid and xylose (group effect, p-value <0.05) whereas over the 30-minute period, the almond group had greater AUC_30min_ for 2-hydroxybutanoic acid and myo-inositol compared to the cracker group (group effect, p-value <0.05). Differential group effects were also observed over the first 15 minutes of the oGTT, with the almond group having lower galactonic acid AUC_15min_ compared to the cracker group (group effect, p-value <0.05).

### Metabolites involved in lipid metabolism

Based on the AUC_120min_ measures, the almond group had lower isoheptadecanoic acid, PE (p-36:1) or PE (o-36:2), PC (42:6), and TG (48:0), and greater 1/2-OG, 11-HETE, 12(13)-EpOME, 9-KODE, 15-HETE, 9,10-DiHODE, C16:1n7t, AA, 9,10-e-DiHO, and C12:0 in comparison to the cracker group (group effect, p-value <0.05). Over the 60-minute period, the almond group also had lower AUC_60min_ for 17,18-DiHETE and 3-hydroxybutyric acid, and greater AUC_60min_ for 12-HETE, 15,16-DiHODE, DG(36:3), C18:1n9, C16:0, LPC (p-18:0) or LPC (o-18:1), C22:6n3, PE(38:4), 9(10)-EpOME, and TXB2 compared to the cracker group (group effect, p-value <0.05). Differential group effects for various lipids were also observed over the first 15- and 30-minute periods (**Figure 4**). Diglyceride and triglyceride clusters are not depicted in the network map for clarity and have been discussed in the previous sections.

### Energy metabolism

The almond group had greater AUC_120 min_ for malic acid, phosphate, and aminomalonate compared to the cracker group (group effect, p-value <0.05, **Figure 4**).

### Metabolites involved in nucleotide metabolism

The almond group had greater AUC_15 min_ for hypoxanthine and inosine, and greater AUC_30 min_ for uridine compared to the cracker group (group effect, p-value <0.05, **Figure 4**).

### Metabolites involved in xenobiotics biodegradation and metabolism

The almond group had greater AUC_120 min_ for 7-methylxanthine compared to the cracker group (group effect, p-value <0.05, **Figure 4**).

### Correlations of metabolites with MI

The PLS-DA results demonstrate distinct clusters separating the almond and cracker groups (**Figure 5**). Thirty-two metabolites (including MI) had loading values within 1.5 SD from the mean loading values. The correlation plots of these metabolites are shown separately for the almond and cracker groups (**Supplementary Figures 1a and 1b**). Importantly, in the almond group, MI demonstrated a moderate negative correlation with CE (22:6) (r =-0.46) and a moderate positive correlation with 12,13-DiHOME (r =0.45) (P <0.05 for almond group; and P <0.05 for almond vs. cracker correlations).

**Figure 5.**
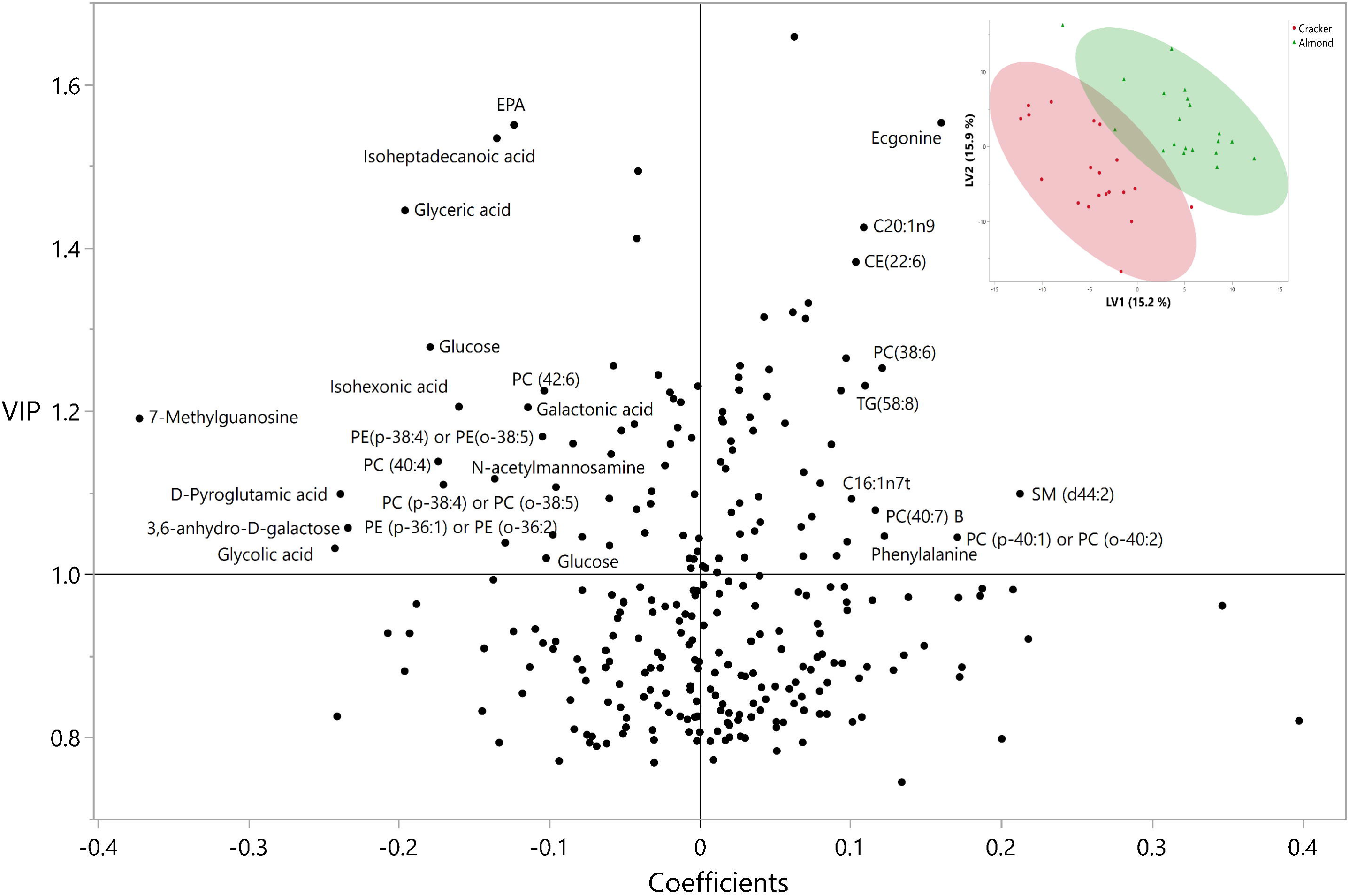
VIP vs. coefficient plot of the metabolites from the PLS-DA analysis conducted on the metabolite AUC and indices to differentiate between almond and cracker groups. Labels only marked for metabolites with VIP ≥ 0.1, and coefficients ≥ 0.1 or ≤ -0.1. Not all data points are shown. Plot of latent variable 1 versus 2 of the PLS-DA analysis inset.

## DISCUSSION

This study characterized changes in the human metabolome following an acute glucose challenge and investigated variations in these responses between individuals consuming almonds versus graham crackers for 8 weeks. As shown previously [9,13–16], a progressive temporal shift in response to a glucose challenge was observed with alterations across amino acid, carbohydrate, and lipid metabolism pathways. Findings also exhibit a notable biphasic lipid response distinguished by higher levels of certain lipids such as unsaturated triglycerides in the earlier periods of a glucose challenge followed by lower levels in the latter period in the almond versus the cracker group. At the individual metabolite level, almond consumption was associated with distinct shifts in total AUC_120 min_ of specific amino acids and lipid mediators involved in metabolic and physiological processes.

The dynamic response of the human metabolome to a glucose challenge as depicted by an oGTT can help reveal metabolic alterations associated with health and disease [17]. The overall lack of significant metabolic changes over the first 15 minutes likely reflects a latent period in which glucose continues to reach peak absorption levels, to saturate perfused tissues, and the delay in the cell’s capacity to respond to an acute increase in extracellular glucose. This is in clear contrast to the latter oGTT time periods, which are characterized by more dynamic shifts in metabolic pathways. These shifts reveal the effects chronic almond consumption have on the cellular adaption to an acute glucose load and activation of specific metabolic pathways over longer durations [17]. The most dynamic metabolic changes have been reported over the first 90 minutes with no difference between the 90 and 120 minute oGTT time points [1]. While most studies have examined changes in metabolomic profiles in the fasted state, studies that have captured multivariate shifts in profiles during an oGTT demonstrate alterations in metabolites of dietary or microbial origin [1], those associated with mitochondrial efficiency and glucose oxidation [1], or markers of proteolysis and lipolysis associated with insulin sensitivity [18].

In the present study, the clear demarcation between the almond and cracker groups in the PLS-DA plots suggests that despite the isocaloric nature of the interventions, the inherent differences in the composition of the two foods, acutely and dynamically influence an array of metabolic pathways. As previously reported, at the end of the 8-week intervention, the almond group’s diet consisted of 42.7% carbohydrates and 41.7% fats, whereas the cracker group consumed 53.7% carbohydrates and 33.5% fats on average. These nutritional differences likely influenced the lipid responses during the glucose challenge, as further illustrated by the MESH class enrichment analysis. Since unsaturated triglycerides were enriched in the almond group in the end-of-study sample following an overnight fast [5], it may be reasonable to assume that the greater levels over the first 30 minutes of the glucose challenge reflect the already elevated circulating levels secreted to help support the short-term fasting metabolism [5]. These differences persist despite adjustment for time 0 values, providing some validation that the changes were induced by chronic almond consumption and reflect the corresponding metabolic adaptations. In contrast, saturated triglycerides were only enriched in the latter periods of the oGTT with lower levels observed in the almond group. The pattern mirrors the lower levels of unsaturated fats in the latter time periods suggesting that the glucose stimulated insulin may have facilitated the clearance of fats. Insulin released in response to the glucose bolus during the oGTT can enhance lipoprotein lipase activity. This increase in lipoprotein lipase activity and that of remnant receptors is essential for the efficient clearance of triglycerides from the bloodstream [19–21]. In this same study, we observed greater insulin sensitivity in the almond group compared to the cracker group [4] suggesting that this improvement in insulin sensitivity may facilitate the observed metabolism of TGs. These interactions suggest that chronic almond consumption selectively impacted the metabolism of saturated triglycerides differentially from unsaturated fats during the early stages of the glucose challenge particularly when comparing the effects to cracker consumption.

At the individual metabolite level, the AUC for isoheptadecanoic acid, a type of saturated fatty acid, was differentially lower in the almond group compared to the cracker group. Heptadecanoic acid, often used as a biomarker for dairy fat intake and linked to a decreased risk of type 2 diabetes, is a related compound [22,23]. High-fiber diets are found to reduce urine levels of this metabolite [24]. Its branched-chain variant, isoheptadecanoic acid, constitutes about 2% of dairy fat and can also be produced from branched chain amino acids through *de novo* synthesis [22,25]. Research involving children has shown an inverse relationship between plasma levels of isoheptadecanoic acid and hepatic steatosis [22]. The reduced AUC for isoheptadecanoic acid in the almond group suggests that chronic almond consumption shifts lipid metabolism from *de novo* lipogenesis to enhanced utilization of fatty acids, which may ameliorate the risk of T2D. The improvement in insulin sensitivity observed in the almond group may facilitate more efficient lipid metabolism, potentially reducing hepatic *de novo* lipogenesis [26] and the risk of hepatic steatosis.

In contrast, almond consumption resulted in a greater AUC for: **a)** AA, an omega-6 fatty acid precursor to eicosanoids; **b**) cyclooxygenase/lipoxygenase-derived hydroxy fatty acids (i.e., 11-HETE, 15-HETE and 12-HETE (p-value=0.092)), and **c)** cytochrome P450/soluble epoxide hydrolase-derived epoxy and dihydroxy fatty acids (i.e. 9,10-e-DiHO; 9,10-DiHODE, 12,13-EpOME and 12,13-DiHOME (p-value=0.089)). The effects of oxylipins on metabolic and physiological processes are nuanced and structurally dependent [27,28]. While 11-HETE and 15- HETE are associated with COX metabolism [29], 15-HETE can also exhibit anti-inflammatory properties [27]. Additionally, while 12-HETE decreases insulin secretion *in vitro* [30], it is also decreased in obesity and inversely associated with arterial stiffness even after BMI adjustment [31]. Little is known about the metabolic impacts of oleic acid and alpha-linolenic acid-derived epoxides and diols, however, the analogous linoleate metabolites have received some attention. For example, previous case studies have documented postprandial changes in 12,13 EpOME following high-carbohydrate challenges (such as consuming a banana) [32]. These epoxides are metabolized by soluble epoxide hydrolase (sEH) into 12,13-DiHOME and its counterpart, 9,10- DiHOME [33,34]. Evidence suggests that the 12,13-DiHOME may contribute to fatty acid uptake and oxidation in adipose tissue and skeletal muscle [33,35]. In addition, a negative correlation of circulating 12,13-DiHOME with insulin resistance has previously been demonstrated [35]. Adding to this evidence base, our study demonstrates a positive correlation of 12,13-DiHOME AUC with postprandial insulin sensitivity in response to chronic almond consumption, suggesting that the improved insulin sensitivity with almond consumption [37] could potentially be mediated via this lipokine. However, it’s important to note that the concentration of circulating 12,13-DiHOME can also be affected by several factors such as exercise and temperature [33].

Correlations between other omega-3 derived metabolites and MI provide additional insights. The negative correlation between MI and CE (22:6) AUC in the almond group suggests that individuals with higher insulin sensitivity may have a suppressed response in CE (22:6) production or release after glucose intake and implicates that those with better insulin sensitivity were more effective at utilizing or clearing it from the bloodstream during the glucose challenge. CE 22:6 is a cholesteryl ester containing DHA, an omega-3 fatty acid [38] and although not statistically significant, the almond group exhibited a trend toward higher CE 22:6 AUC compared to the cracker group (P =0.051). CE 22:6 has not been widely studied, and those that have, reported this metabolite in the pathogenesis of osteoarthritis, Parkinson’s disease, and bladder cancer [39–41]. However, it appears that in healthy individuals the incorporation of DHA in LDL cholesterol esters does not significantly promote LDL oxidation – a factor implicated in atherosclerosis [42]. Moreover, a systematic review reported that DHA intake can lower oxidative stress [43]. It’s essential to juxtapose this with the observation of reduced omega-3 total fatty acids (TFAs) in the almond group in the fasting state [5], which contrasts with the more dynamic lipid profile seen during the oGTT. Their reduced levels following an overnight fast in the almond group may indicate potential interactions between different unsaturated fatty acids, the intricate metabolism involving a series of steps [44], or possibly the overriding influence of other bioactive compounds in almonds.

Almond consumption also induced lower AUC for L-cystine compared to the cracker group. L-cystine is an oxidized dimer of L-cysteine which is a precursor for the synthesis of glutathione, a potent, non-enzymatic antioxidant [45]. Given that there were no significant differences in cysteine or L-cysteine-glutathione disulfide AUC between groups, and considering glutathione’s role in enhancing cellular redox balance and mitigating oxidative stress, the lower L-cystine AUC could instead indicate a more regulated or less stressed oxidative state in the almond-consuming individuals [46,47]. Since plasma glutathione levels are usually relatively low in normal, healthy humans [48] and glutathione synthesis is activated by acute increases in free radicals [49], the lower AUC for L-cystine without a change in cysteine, could indicate that chronic almond consumption helps maintains a balanced redox state through the efficient use of cysteine reserves during post-prandial, hyperglycemic excursions [50,51]. The lower L-cystine AUC may also reflect an environment where there’s less necessity to combat oxidative stress, possibly due to the inherent antioxidant properties of almonds [52,53]. This state could contribute to better cardiovascular health by maintaining efficient glutathione induced regulation of nitric oxide [46,47], aligning with the cardiovascular benefits associated with almond intake [54].

Almond consumption also resulted in lower AUC for N-acetylmannosamine, which is an important precursor in the biosynthesis of sialic acids [55]. Sialic acids are important components of glycoproteins and glycolipids and critically important in cellular communication and signaling [56,57]. The lower AUC of N-acetylmannosamine might reflect reduced synthesis and/or more efficient utilization of sialic acid-containing glycans. Notably, our previous analyses of fasting data revealed a positive correlation between fasting serum N-acetylmannosamine levels and the predicted microbial community potential to produce this metabolite in the almond group [5]. Given that the gut microbiome has been shown to predict postprandial glucose metabolism [58,59], it is plausible that a chronic alteration in gut microbiota as demonstrated in our study [5,60], could impact the synthesis and degradation of glycan-related compounds during metabolic challenges such as an oGTT.

### Summary

The study underscores the dynamic nature of the human metabolome in response to a glucose challenge following chronic almond consumption when compared to an isocaloric cracker control. Our findings demonstrate a distinctive biphasic lipid response to an acute glucose load characterized by differentially greater levels of unsaturated triglycerides in the initial phases of the glucose challenge, followed by lower levels in the later stages in the almond group compared to the cracker group. The acute glucose challenge also revealed significant alterations across amino acid and lipid mediators involved in metabolic and physiological pathways that had not been explored previously. Study findings also highlight the intricate interactions between diet, metabolome, and insulin sensitivity. Future research should delve deeper into the specific pathways responsible for the observed metabolic changes using kinetic analyses, particularly the mechanisms driving differential lipid and fatty acid responses in the almond group. The differences between the glucose challenge (acute, dynamic) and an overnight fasting (chronic, static) results [5] accentuate the need to view dietary impacts through both acute and chronic lenses, as both offer unique insights into the role of diet in shaping the metabolic determinants of health and disease.

## AUTHOR CONTRIBUTIONS

RMO and JD conducted the study. JWN and OF directed the metabolomics analyses. JD and SP drafted the manuscript. All authors revised the manuscript and approved the submitted version.

## DATA AVAILABILITY

Data is available at https://doi.org/10.6084/m9.figshare.17132201

