## Supplemental Figure 1 for "Metabolic Responses to an Acute Glucose Challenge: The Differential Effects of Eight Weeks of Almond vs. Cracker Consumption in Young Adults"

**Supplementary Figure 1**. Heat map of Pearson’s correlations of metabolite fasting and AUC 120 variables in a) the almond and b) cracker groups. In the extremes of the color gradient, red represents a strong positive correlation while blue represents a strong negative correlation. The size of the correlation represents the statistical significance i.e., big rectangle indicates P<0.05, small rectangles indicate P>0.05, missing rectangles indicate that correlations are between the same variables.

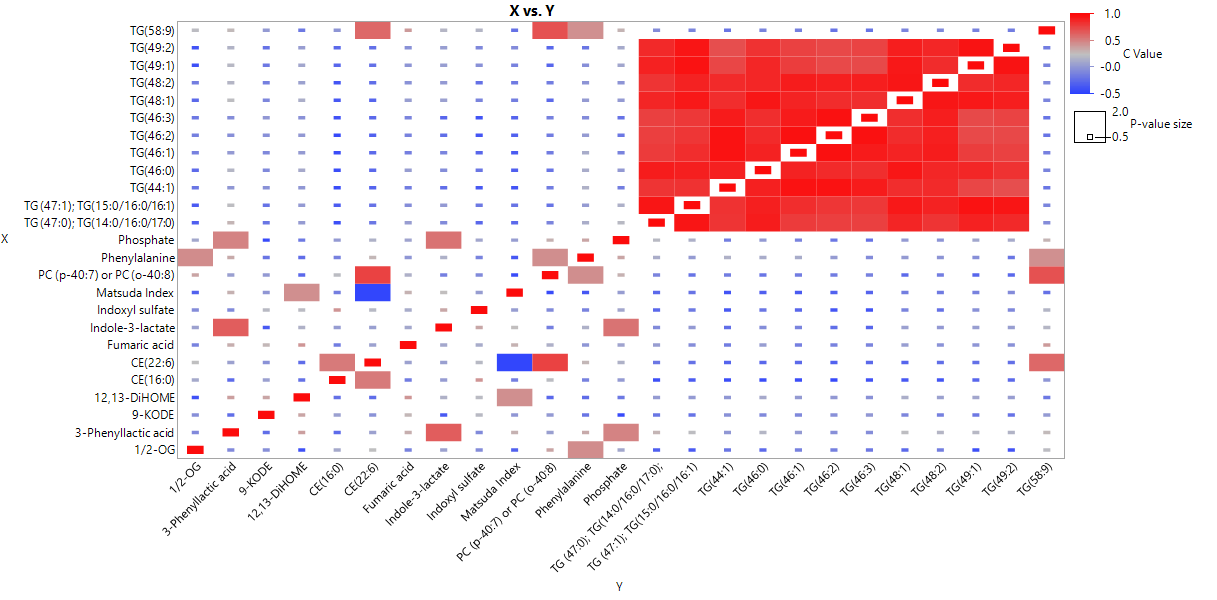

**A)**

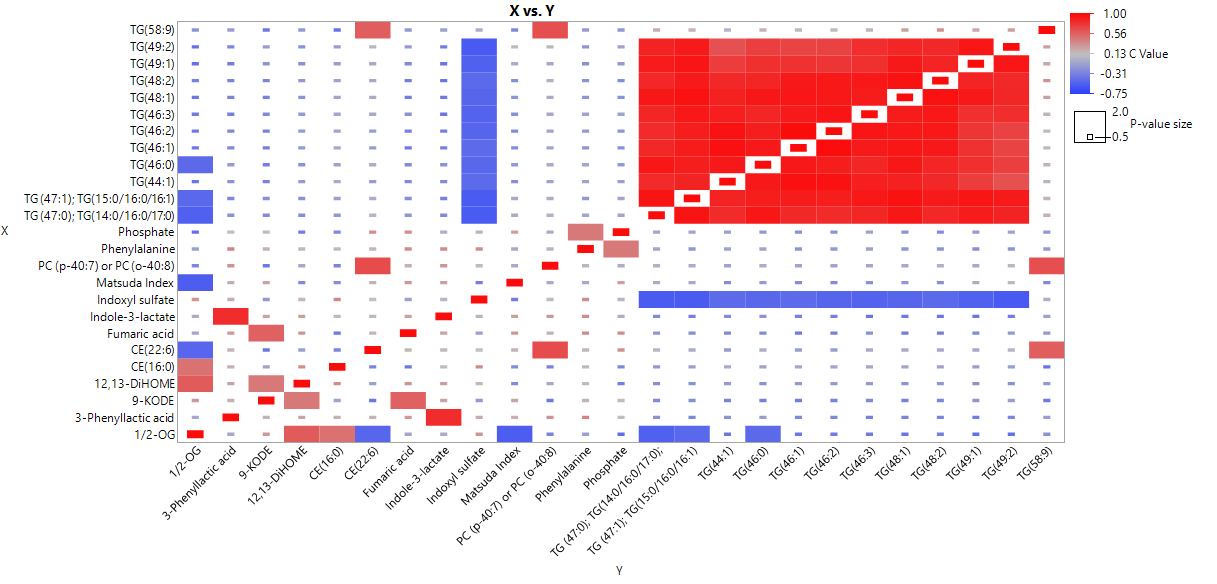

**B)**
